# Association Between Beta Blocker and Clinical Outcome in Adult Patients with Sepsis or Septic Shock: Protocol of a Systematic Review and Meta- Analysis of Randomized Controlled Trials

**DOI:** 10.1101/2023.11.14.23298548

**Authors:** Sulagna Bhattacharjee, Emmanuel Easterson Ernest, Souvik Maitra

## Abstract

Use of beta-blockers as a part of heart rate control strategy is sepsis and septic shock patients is of great debate. Despite of early encouraging results, no large trial was performed and several subsequent small studies reported conflicting results. This meta-analysis and systematic review will be conducted and published as per PRISMA guidelines. In this review, randomized controlled trials comparing short-acting beta-blockers with ‘standard of care’ in adult patients with sepsis and septic shock will be included. Primary outcome will be 28-day mortality and secondary outcomes will be duration of intensive care unit stay, duration of hospital stay, ICU mortality, hospital mortality and reported adverse events. A random effect model will be used for all analysis.

**Source of Support:** Nil

**Conflict of interests:** None

## Introduction

Sepsis is a common cause of hospital admission and the commonest cause of intensive care unit (ICU) admission worldwide [1]. The estimated case burden of sepsis in India was 11 million per year, and nearly one-third of them died [2]. Though worldwide reported mortality from sepsis is variable, it may be as high as 40%. Sepsis carries not only very high mortality; it causes significant disability in the survivors. Nearly 30% of the patients who survive initial insult have a chronic protracted course with multiple episodes of hospital-acquired infection, organ dysfunction, and disability, causing prolonged ICU stay [3].

Activation of adrenergic sympathetic system in the early phase of sepsis is almost universal and possibly associated with several adverse clinical effects [4]. Persistent tachycardia may be associated with increased myocardial demand, shorter duration of diastole and associated with poor outcome [5]. Pre-clinical and some clinical studies reported encouraging results with the use of both short acting and long acting beta-blocker in septic shock patients [6, 7]. However, recently, a well-designed randomized controlled trial [8] reported contradictory findings making this systematic review and meta-analysis necessary.

## Methods

PRISMA (Preferred Reporting Items for Systematic Reviews and Meta-Analyses) guidelines will be followed for conduction and reporting of this study [9]. Protocol of this systematic review and meta-analysis will be registered/ published in a publicly accessible database.

### Eligibility Criteria

This systematic review and meta-analysis will include randomized control trial comparing beta-blocker with ‘standard of care’ in adult patients with sepsis and septic shock. Studies using either short acting or long acting beta-blockers will be eligible for inclusion. Observational studies, retrospective studies, case series and reports will not be eligible for inclusion.

### Information Sources

PubMed, Scopus and Google scholar database will be searched manually from inception to 15^th^ November 2023 for identification of potentially eligible studies. No language restriction will be applied in the search strategy.

### Search Strategy

PubMed will be searched with the following search words: ‘beta blocker and sepsis’, ‘sepsis and esmolol’, ‘sepsis and landiolol’, ‘septic shock and landiolol’, ‘short acting beta blocker and sepsis’, ‘beta blocker and septic shock’, ‘esmolol and septic shock’, ‘propranolol and sepsis’, ‘propranolol and septic shock’, ‘metoprolol and sepsis’, ‘metoprolol and septic shock’, ‘sepsis and heart rate control’, ‘septic shock and heart rate control’. Similar search words will be used for both Scopus and Google scholar also. Duplicate publications will be removed electronically by Endnote software. Detailed search strategy will be reported in the appendix of the study.

### Selection Process

All unique articles will be assessed from title and abstract for inclusion in this study. Two study authors will assess articles independently and in case of any dispute between the assessors, opinion of the third author will be considered as final. Full texts of the potentially eligible studies will also be assessed by two independent study authors for final selection and opinion from the third author will be taken in case of any disagreement.

### Data Collection Process

Required data will be retrieved from the full text and supplementary content (if applicable) of the included trials. All data will be collected independently by two study authors and also cross-checked by the other author for any discrepancy.

### Data Items

Following study details will be retrieved: Year of publication, name of first author, country(s) where study was conducted, sample size, inclusion and exclusion criteria of the participants, baseline characteristics of the study participants, drug, dosage and duration of the beta-blocker therapy, details of the standard of care treatment and duration of follow-up. Primary outcome of this meta-analysis will be reported 28-day mortality. Secondary outcomes will be duration of intensive care unit stay, duration of hospital stay, ICU mortality, hospital mortality and reported adverse events.

### Risk of Bias Assessment

Risk of biases will be assessed as per Cochrane methodology (Risk of Bias tool version 2) by two independent study authors. The following domains will be assessed from each study: randomization process, deviation from the intended intervention, missing outcome data, bias in measurement of outcome and bias from selection of reported outcome.

### Effect measures and Data Synthesis Methods

All extracted data will be tabulated in a spreadsheet (Microsoft Excel datasheet). For a continuous outcome, mean and standard deviation (SD) values will be tabulated from both arms of the study, a mean difference (MD) will be calculated at the study level, and a weighted mean difference will be calculated to pool the results across all trials. If the values were reported as median and an inter-quartile range or total range of values, the mean value will be estimated from a previously described method [10]. Risk ratio (RR) for each trial and pooled RR using the inverse variance method will be computed for binary variables. All statistical variables will be calculated with 95% confidence interval (95% CI). The Q-test was used to analyze heterogeneity of trials. Considering possible heterogeneity due to study design and patients’ population, we used a random effect model for all pooled analysis. Pooled analysis will be conducted in RevMan software (Review Manager (RevMan) [Computer program]. Version 5.3. Copenhagen: The Nordic Cochrane Centre, The Cochrane Collaboration, 2014). Publication bias will be tested by Egger’s regression test.

## Results

Results of this systematic review and meta-analysis will be reported as per PRISMA guidelines and will be published in a peer reviewed journal.

## Data Availability

Not applicable

